# How deep brain stimulation and levodopa affect gait variability in Parkinson’s disease

**DOI:** 10.1101/2020.10.07.20207704

**Authors:** Zi Su, Salil Patel, Bronwyn Gavin, Tim Buchanan, Marko Bogdanovic, Nagaraja Sarangmat, Alexander L. Green, Tipu Z. Aziz, James J. FitzGerald, Chrystalina A. Antoniades

## Abstract

**Background:** Disorders of gait are a very common feature of Parkinson’s Disease. We examined how deep brain stimulation of the subthalamic nucleus (STN DBS) and dopaminergic medication affect gait and more specifically its rhythmicity.

**Objectives:** We accurately quantified multiple gait parameters in Parkinson’s patients during on and off stages of their treatment (levodopa or STN DBS) to compare and contrast the treatment-induced changes in gait.

**Methods:** We studied 11 patients with STN DBS, 15 patients on levodopa and 42 healthy controls. They all completed the MDS-UPDRS part III along with a gait assessment protocol while wearing six nine-axis inertial measurement units (lumbar, sternal, and all four extremities).

**Results:** Both medication and stimulation significantly improved stride length, while medication further significantly increased gait speed. In the lower limbs, both medication and stimulation had a normalising effect on lower limb angles, significantly increasing the foot strike angle and toe-off angle.

**Conclusions:** STN DBS reduced the step to step variability in a range of lower limb gait parameters in PD, while antiparkinsonian medication had no significant effect. This suggests that STN stimulation, but not dopaminergic medication, has access to circuits that control gait rhythm, and that the resulting effect of stimulation on gait is beneficial. However, the results we observed for movement of the trunk and upper limbs were strikingly different to those seen in the lower limbs. We propose a hypothesis to explain why we observe these results, focusing on cholinergic pedunculopontine projections.

## Introduction

Disorders of gait are a very common feature of Parkinson’s disease (PD). They can be classified into two types, which commonly coexist: gait speed and amplitude abnormalities due to bradykinesia and rigidity, and gait rhythm disturbances which in their most severe form manifest as gait freezing. Impaired gait significantly affects the quality of life of PD patients and increases the risk of falls (1). One aspect of gait that is of particular importance is its regularity, or lack thereof. There is a clear association of gait irregularity with falls risk (1, 2), and a reduction in gait irregularity may therefore have a direct effect in preventing falls and consequent injury.

Overall straight-line gait speed, and the amplitude of parameters including step length, trunk motion and arm swing, are improved by dopaminergic medication (3). Other parameters such as cadence, turning in place, and aspects of gait initiation are less responsive (4). Published work is suggestive of decreased gait variability with medication (1, 3), but the literature is sparse.

Deep Brain Stimulation (DBS) of the subthalamic nucleus (STN) is an effective treatment for PD, improving rigidity, bradykinesia, and tremor, and reducing motor fluctuations (5, 6). It also usually results in a substantial reduction in levodopa requirements, which may in turn reduce levodopa-induced side effects. DBS improves many of the same gait parameters as medication, including overall speed and stride amplitude (7). A single previous study has suggested that STN DBS reduces parkinsonian gait variability (8).

In this study we examine in detail how STN DBS and dopaminergic medication affect gait and specifically its variability. We used a wearable sensor array to accurately quantify multiple gait parameters in PD patients both on and off their treatment (levodopa or STN DBS) to compare and contrast the treatment-induced changes in gait. Often in previous studies the variability present within each parameter has been expressed in terms of its standard deviation or coefficient of variation for multiple gait cycles in a testing session. However, these measures potentially conflate long term variability (e.g. gradual slowing) with short term variability (true irregularity); we are primarily interested in the latter and have used analysis methodology that aims to separate this out.

Measuring gait variability has become much easier in recent years with the proliferation of wearable inertial measurement devices (9) that are replacing motion detection camera systems, video recording analysis, and force sensitive mats in many gait laboratories. While studies conducted previously often analysed only a few measures, modern systems can report on in excess of 100 variables. Which ones are the most clinically relevant is an important and unexplored question. Here we have examined a set of 18 gait parameters that are prevalent in the literature.

## Methods

### Participants

Participants were recruited through the OxQUIP (Oxford Quantification in Parkinsonism) study at the John Radcliffe Hospital in Oxford. OxQUIP is a large prospective cohort study of neurophysiological biomarkers in parkinsonian patients and is approved by the ethics committee (REC reference 16/SW/0262). Informed consent was obtained from all participants after explaining the procedures to them. PD participants had a diagnosis of idiopathic PD according to UK PD brain bank criteria (10). Age matched healthy controls were often but not always the spouse of a patient.

Two groups of PD patients were recruited. A group with STN DBS systems were tested with stimulation on and then again with stimulation off. Another group of patients without DBS systems but at a similar disease stage were tested in the medication off and medication on states. The medication off state is defined as at least 12 hours without antiparkinsonian medication, and the medication on state is one hour post dose. Age-matched healthy controls were tested once.

### Gait testing

Testing consisted of gait analysis and MDS-UPDRS (Movement Disorder Society Unified Parkinson’s Disease Rating Scale) part III (motor assessment). Gait data were collected using the APDM MobilityLab System (http://www.apdm.com/mobility) comprising a network of six synchronized, body worn inertial measurement units (triaxial accelerometers, gyroscopes, and magnetometers) positioned on both wrists and feet, the trunk, and the lumbar area. Patients performed a two-minute walk in a quiet, uncarpeted 15m corridor making turns when necessary.

The parameters analysed are shown in figure 1 and table 2 (in supplemental materials) and included a range of lower limb, trunk, and upper limb measures. Values for each subject are the mean of right and left sided values.

**Figure 1.**
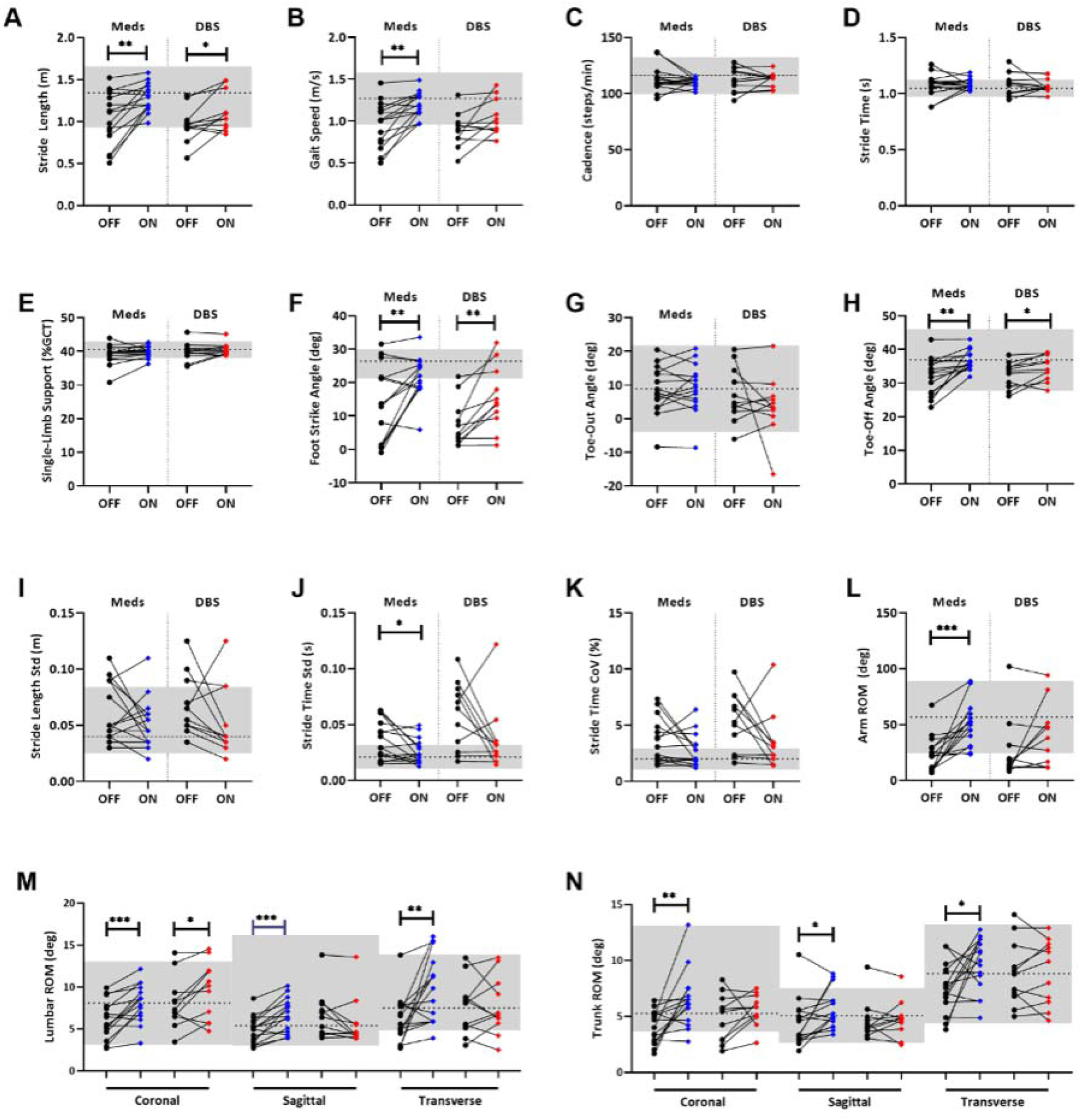
Treatment effect of STN DBS and medication on selected gait parameters against the normative range. Black, off treatment; Blue, on medication; Red, on DBS; Grey, normative range, calculated as the 95% confidence interval of each parameter from the age-matched control group; SD, standard deviation; ROM, range of motion; CoV, covariance. * denotes *p* < 0.05, ** denotes *p* < 0.01, and *** denotes p < 0.001.

### Analysis

Step by step measures of gait parameters were exported from MobilityLab and processed using GraphPad Prism (GraphPad Software, www.graphpad.com). Poincaré analysis was used for quantification of variability during gait. A Poincaré plot, also known as a first return map, is a scatter plot in which each successive value of a given variable (for example stride length) is plotted against the one before, i.e. *x* = *f*(*i*); *y* = *f*(*i*+1). This is shown schematically at the top left of figure 3. Deviation of points from the line of identify (*y* = *x*) occurs due to step-to-step variability in the parameter concerned. This short-term variability is quantified using the standard deviation of the distance from the identity line, known as SD1, where:

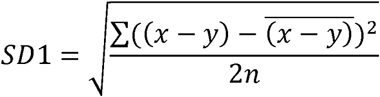

The spread of data points along the identity line represents long term variability, for example due to gradual slowing due to fatigue over the testing session. It is quantified by its standard deviation SD2, where:

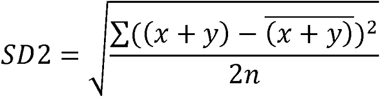

SD1 and SD2 were calculated for each parameter analysed for each participant in each condition.

Groups were compared using paired *t*-tests where data were normally distributed and the Wilcoxon signed rank test (WSRT) otherwise.

## Results

Demographic data, clinical characteristics, and rating scores are shown in Table 1. For simplicity within this paper, participants without DBS, who are tested on and off medication, are referred to as the medication group, and those with DBS, who are tested on and off stimulation, are referred to as the DBS group. DBS patients were tested in the on medication condition throughout; STN DBS typically reduces medication requirements by 30-50% and in this case the DBS group had a mean dose in levodopa equivalent units (LEU) that was 42% less than the medication group.

**Table 1.** Demographics, clinical characteristics and rating scale scores in both the PD patient and healthy control groups. ID, identification number; PD, Parkinson’s disease; MDS-UPDRS, Movement Disorders Society Unified Parkinson’s Disease Rating Scale; H&Y ON, Hoehn and Yahr score on treatment; H&Y OFF, Hoehn and Yahr score off treatment; DBS, Deep Brain Stimulation; LEU, levodopa equivalent unit; M, male; F, female. Figures in parantheses are ranges.

| ID | Age (yrs) | Gender | PD Duration (yrs) | MDS-UPDRS PART 3 ON | MDS-UPDRS PART 3 OFF | H&Y ON | H&Y OFF | DBS duration (yrs) | LEU (mg) | Medication |
| --- | --- | --- | --- | --- | --- | --- | --- | --- | --- | --- |
| DBS01 | 51-65 | F | 18 | 33 | 40 | 3 | 3 | 4 | 1235 | Amantadine, Co-beneldopa, Co-careldopa CR, Entacapone/levodopa/carbidopa, Ropinirole |
| DBS02 | 51-65 | F | 15 | 29 | 36 | 2 | 2 | 2 | 410 | Co-beneldopa, Rasagiline, Ropinirole |
| DBS03 | 51-65 | M | 10 | 34 | 93 | 0 | 3 | 1 | 700 | Amantidine, Co-careldopa, Entacapone/levodopa/carbidopa |
| DBS04 | 51-65 | M | 7 | 26 | 38 | 1 | 1 | 3 | 400 | Co-beneldopa |
| DBS05 | 51-65 | M | 12 | 36 | 50 | 2 | 3 | 2 | 250 | Amantadine, Co-careldopa |
| DBS06 | 51-65 | M | 12 | 60 | 72 | 2 | 2 | 5 | 650 | Co-beneldopa, Pramipexole PR |
| DBS07 | 51-65 | M | 10 | 37 | 58 | 2 | 2 | 3 | 350 | Amantadine, Co-careldopa |
| DBS08 | 51-65 | M | 11 | 34 | 55 | 2 | 2 | 3 | 620 | Co-careldopa, Rotigotin |
| DBS09 | 51-65 | F | 3 | 40 | 68 | 3 | 3 | 2 | 0 | None |
| DBS10 | 51-65 | F | 11 | 28 | 38 | 2 | 2 | 2 | 200 | Co-careldopa, Pramipexole PR |
| DBS11 | 51-65 | M | 9 | 17 | 48 | 2 | 2 | 1 | 435 | Co-careldopa, Pramipexole PR, Rasagiline, Opicapone |
| **Stimulation group** | 58.9±4.4 (51-65) | 7M:4F | 10.7±3.7 (3-18) | 34.0±10.2 (17-60) | 54.2±16.9 (36-93) | 1.9±0.8 (0-3) | 2.3±0.6 (1-3) | 2.5±1.1 (1-5) | 477±310 (0-1235) |  |
| MED01 | 54-74 | M | 8 | 18 | 41 | 2 | 2 |  | 1564 | Co-beneldopa, Co-careldopa, Entacapone, Rasagiline |
| MED02 | 54-74 | M | 3 | 17 | 29 | 2 | 2 |  | 680 | Co-beneldopa, Ropinirole |
| MED03 | 54-74 | M | 6 | 27 | 49 | 2 | 2 |  | 500 | Co-beneldopa, Rasagiline |
| MED04 | 54-74 | F | 9 | 34 | 42 | 2 | 2 |  | 315 | Apomorphine, Co-careldopa CR, Rotigotine |
| MED05 | 54-74 | M | 3 | 14 | 39 | 2 | 2 |  | 948 | Co-careldopa, entacapone |
| MED06 | 54-74 | M | 7 | 19 | 43 | 2 | 2 |  | 940 | Cobeneldopa MR, Entacapone/levodopa/carbidopa |
| MED07 | 54-74 | F | 12 | 28 | 65 | 3 | 3 |  | 1200 | Co-beneldopa, Entacapone/levodopa/carbidopa |
| MED08 | 54-74 | M | 20 | 39 | 38 | 2 | 2 |  | 300 | Co-beneldopa |
| MED09 | 54-74 | M | 7 | 11 | 44 | 2 | 2 |  | 1140 | Co-beneldopa, Co-beneldopa CR, Pramipexole, Safinamide |
| MED10 | 54-74 | M | 16 | 24 | 59 | 2 | 2 |  | 660 | Apomorphine pen, Co-beneldopa, Opicapone, Rotigotine, Safinamide |
| MED11 | 54-74 | M | 8 | 32 | 41 | 2 | 2 |  | 610 | Co-careldopa, Rasagiline, Ropinirole XL, Trihexyphenidyl |
| MED12 | 54-74 | M | 8 | 27 | 47 | 2 | 2 |  | 1340 | Co-careldopa, Co-careldopa CR, Opicapone, Ropinirole |
| MED13 | 54-74 | M | 7 | 16 | 41 |  | 2 |  | 325 | Entacapone/levodopa/carbidopa, Rasagiline |
| MED14 | 54-74 | F | 5 | 11 | 42 | 2 | 2 |  | 1278 | Co-careldopa CR, Entacapone/levodopa/varbidopa, Rasagiline, Ropinerole XL |
| MED15 | 54-74 | M | 13 | 22 | 58 | 2 | 2 |  | 441 | Co-beneldopa, Co-beneldopa CR, Entacapone, levodopa/carbidopa |
| **Medication group** | 64.9±5.8 (54-74) | 12M:3F | 8.8±4.5 (3-20) | 22.6±8.2 (11-39) | 45.2±8.9 (29-65) | 2.1±0.2 (2-3) | 2.1±0.2 (2-3) |  | 816±401 (300-1564) |  |
| **Control group** | 57.9±6.5 (51-80) | 19M:23F |  | 3.5±4.2 (0-16) |  |  |  |  |  |  |

The effects of medication and DBS on gait parameters are depicted in Figure 1 which shows off and on values of each parameter for the two interventions; each pair of points linked by a line represents a single patient. Gait parameters from the healthy controls were used to generate age-matched normative ranges, defined as the central 95% of the range of control values. These are shown in the figures as grey bands; the horizontal dashed lines are control means. Table 2 (in supplemental materials) gives mean values per parameter, group, and condition, and details of the statistical comparison of off/on values.

**Table 2.** Effect of medication and DBS on gait parameters quantified as mean values per parameter, group, and condition, expressed in parentheses as a percentage of healthy control mean values. HC, healthy control; DBS, Deep Brain Stimulation; pT, Paired-T test; W, Wilcoxon signed rank test; GCT, gait cycle time; deg, degree; Std, standard deviation; CoV, co-variance; ROM, range of motion.

| Parameter | HC group | Medication group |  |  |  | DBS group |  |  |  |
| --- | --- | --- | --- | --- | --- | --- | --- | --- | --- |
|  | Mean | Mean value (% of HC) |  | Significance of change |  | Mean value (% of HC) |  | Significance of change |  |
|  |  | off | on | test | p | off | on | test | p |
| Lower limb |  |  |  |  |  |  |  |  |  |
| Stride length (m) | 1.34 | 1.04 (77) | 1.29 (95) | pT | 0.0016 | 0.964 (72) | 1.11 (83) | pT | 0.019 |
| Gait Speed (m/s) | 1.27 | 0.964 (76) | 1.19 (93) | pT | 0.0012 | 0.895 (71) | 1.04 (82) | pT | 0.051 |
| Cadence (steps/min) | 116 | 113 (97) | 110 (95) | W | 0.45 | 111 (96) | 113 (97) | pT | 0.69 |
| Stride Time (s) | 1.05 | 1.08 (103) | 1.09 (104) | W | 0.80 | 1.09 (104) | 1.07 (103) | W | 0.70 |
| Single-Limb Support (%GCT) | 40.5 | 39.1 (97) | 39.9 (98) | pT | 0.17 | 39.9 (98) | 40.5 (100) | pT | 0.20 |
| Foot Strike Angle (deg) | 26.5 | 13.6 (51) | 21.5 (81) | pT | 0.0062 | 7.72 (29) | 15.4 (58) | pT | 0.0051 |
| Toe-Out Angle (deg) | 8.90 | 8.45 (95) | 9.15 (103) | pT | 0.53 | 7.15 (80) | 3.70 (42) | pT | 0.34 |
| Toe-Off Angle (deg) | 36.9 | 40.0 (87) | 36.7 (99) | pT | 0.0012 | 32.5 (88) | 34.5 (93) | pT | 0.024 |
| Stride Length Std (m) | 0.0400 | 0.0623 (155) | 0.0517 (129) | W | 0.18 | 0.0682 (170) | 0.0532 (133) | pT | 0.19 |
| Stride Time Std (s) | 0.0210 | 0.0358 (170) | 0.0259 (123) | pT | 0.025 | 0.058 (276) | 0.0371 (177) | W | 0.083 |
| Stride Time CoV (%) | 1.99 | 3.56 (179) | 2.75 (138) | W | 0.19 | 5.33 (267) | 3.51 (176) | W | 0.10 |
| Axial |  |  |  |  |  |  |  |  |  |
| Lumbar coronal ROM | 8.09 | 6.12 (76) | 8.13 (100) | pT | 0.0005 | 8.10 (100) | 9.65 (119) | pT | 0.036 |
| Lumbar sagittal ROM | 5.41 | 4.94 (91) | 6.66 (123) | pT | 0.0002 | 6.65 (123) | 5.70 (105) | W | 0.10 |
| Lumbar transverse ROM | 9.40 | 7.70 (82) | 12.2 (130) | pT | 0.0067 | 9.39 (100) | 9.65 (103) | pT | 0.80 |
| Trunk coronal ROM | 5.27 | 4.03 (76) | 6.52 (124) | W | 0.0034 | 4.87 (92) | 5.60 (106) | pT | 0.24 |
| Trunk sagittal ROM | 5.05 | 4.40 (87) | 5.61 (111) | W | 0.025 | 4.62 (91) | 4.75 (94) | W | 0.70 |
| Trunk transverse ROM | 4.38 | 7.57 (173) | 9.61 (220) | pT | 0.030 | 8.86 (202) | 9.00 (205) | pT | 0.78 |
| Upper limb |  |  |  |  |  |  |  |  |  |
| Arm ROM (deg) | 56.6 | 23.3 (41) | 49.8 (88) | pT | 0.0005 | 26.3 (47) | 39.8 (70) | W | 0.41 |

### Lower limb gait parameters

Both medication and DBS significantly improve stride length (Fig. 1A, medication *p* = 0.0016, *t*-test; DBS *p* = 0.019, *t*-test) towards the control mean, while medication further significantly increase gait speed (Fig. 1B, *p* = 0.0012, *t*-test). Medication and DBS also both had a normalising effect on lower limb angles, significantly increasing the foot strike angle (Fig. 1F, medication *p* = 0.0062, *t*-test; DBS *p* = 0.0051, *t*-test) and toe-off angle (Fig. 1H, medication *p* = 0.0012, *t*-test; DBS *p* = 0.019, WSRT). There was no significant treatment effect for absolute toe-out angle. The stride time (or gait cycle duration) was not significantly affected by treatment, however medication did have a significant effect on the standard deviation of stride time (Fig. 1J, *p* = 0.025, *t*-test).

### Axial and upper limb gait parameters

Medication consistently and significantly increases all axial and upper limb range of motion (ROM) parameters, including the arm ROM (Fig. 1L, *p* = 0.0005, *t*-test), lumbar ROM in the coronal, sagittal, and transverse planes (Fig. 1M, *p* = 0.0005, *t*-test; *p* = 0.0002, *t*-test; *p* = 0.0067, *t*-test), and trunk ROM in the coronal, sagittal, and transverse planes (Fig. 1N, *p* = 0.0034, WSRT; *p* = 0.025, WSRT; *p* = 0.030, *t*-test). No such effect is seen with stimulation except for an increase in lumbar coronal ROM of borderline significance (Fig. 1M, *p* = 0.036, *t*-test).

### Variability in gait parameters

Figure 2 (in supplemental materials) shows examples of the step by step values of stride length on and off therapy for the first 5 participants in the medication and DBS groups. In these examples, some of the medication group patients can be seen to have increased variability on treatment (MED01 and MED03), while DBS patients DBS02, DBS03, DBS05 appear to have decreased variability on treatment.

**Figure 2.**
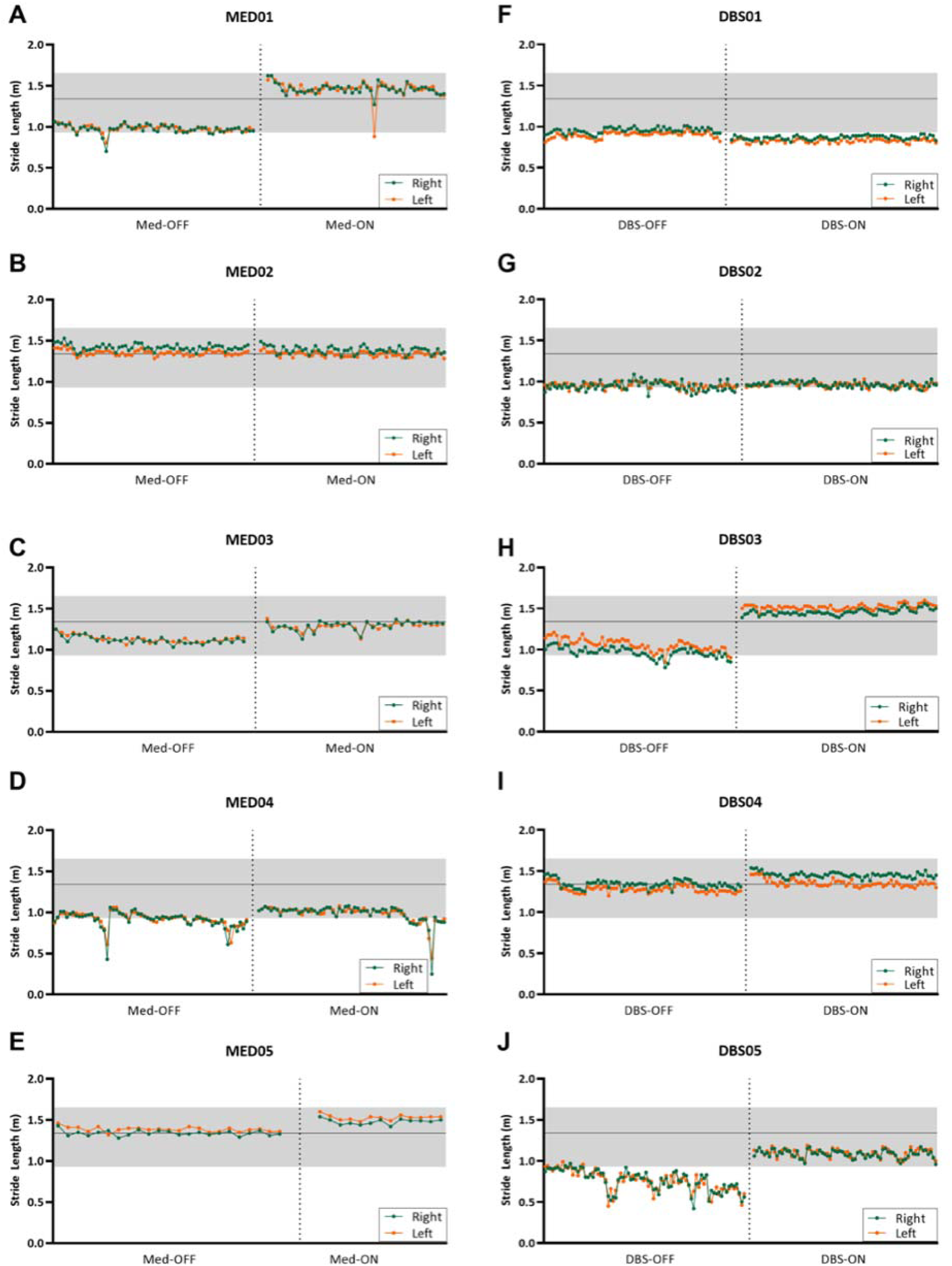
Step-by-step analysis of patient stride length under STN DBS vs medication treatment. Green, right leg; Orange, left leg, Grey, normative range, calculated as the 95% confidence interval of each parameter from the age-matched control group.

To quantify the degree of variability, Poincaré analysis was carried out for each parameter, patient, and treatment condition. As an example, the analysis for toe out variability is shown in figure 3. The larger plot at top left shows the principles of Poincaré analysis, with the standard deviation perpendicular to (SD1) and along (SD2) the identity line being calculated separately, and respectively representing short- and long-term variability. Data from a sample of healthy controls is shown in the top two rows. In rows 3-5 data from the medication group is shown; the scatter of points is in many cases greater in the on medication condition (blue points) than the off medication condition (black points), signifying an increase in variability. The lower two rows are the DBS group; the points in the on DBS condition (red points) are in many cases clustered more tightly than in the off DBS condition (black points), signifying decreased variability.

**Figure 3.**
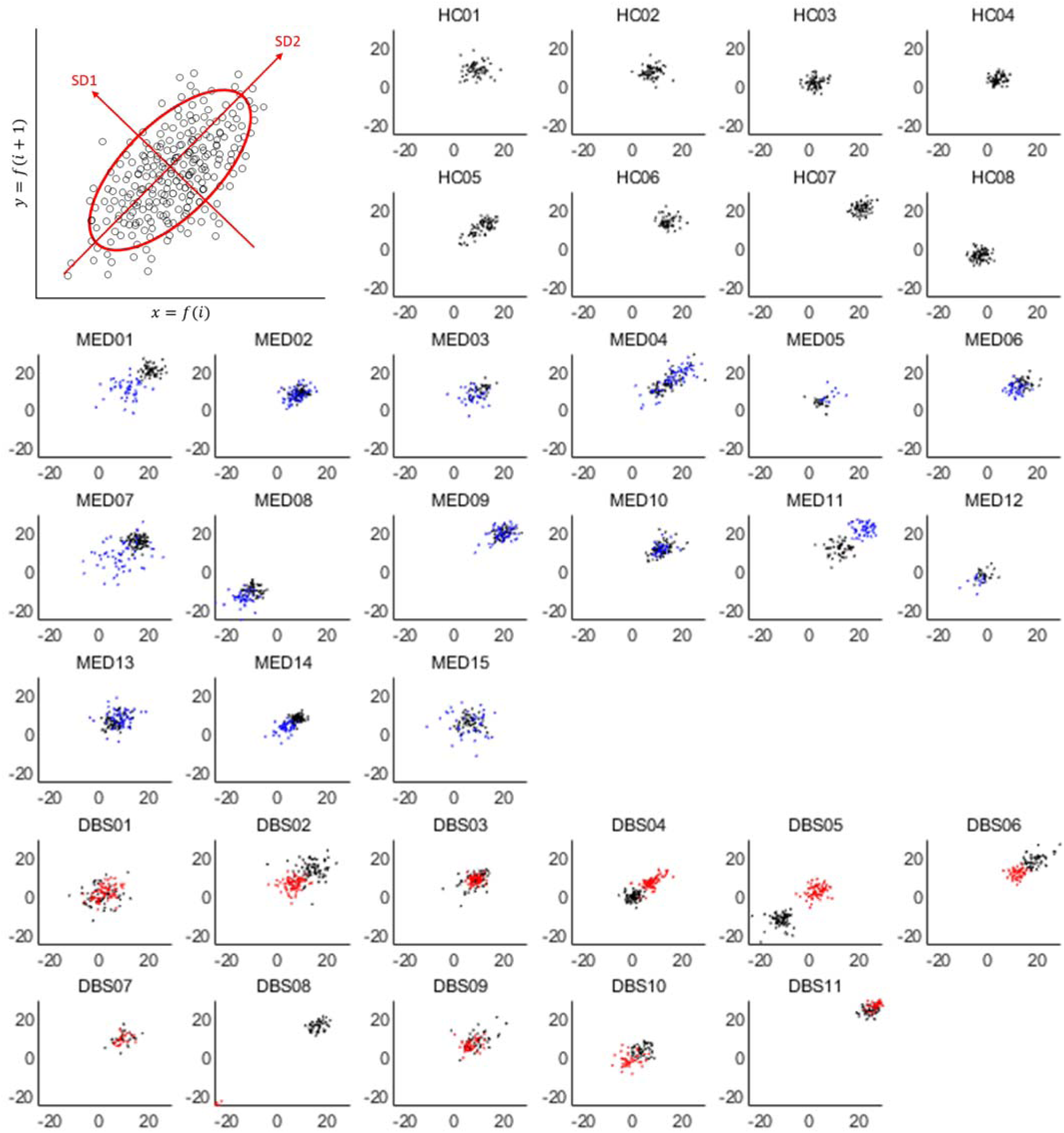
Poincaré plots of toe out angle on and off treatment in the STN DBS and medication groups and the control group. Larger plot at top left shows the principles of Poincaré analysis, with the standard deviation perpendicular to (SD1) and along (SD2) the identity line being calculated separately, and respectively representing short-and long-term variability. The upper two rows of small graphs show data from a sample of the healthy controls. The third to fifth rows are the medication group and sixth and seventh rows are the DBS group. Black, off treatment; Blue, on medication; Red, on DBS. Axis scale unit in degrees.

Figure 4 shows the changes in SD1 (short-term variability) and SD2 (long-term variability) produced by medication or DBS. Each point on each plot represents the difference in on versus off treatment variability for an individual patient. In the lower limbs, medication significantly increases short- and long-term toe out angle variability (Fig. 4B, *p* = 0.0015, WSRT; *p* = 0.022, WSRT), while leaving the other parameters unaffected. In contrast, STN DBS decreases both short- and long-term toe out angle variability (Fig. 4B, *p* = 0.021, *t*-test; *p* = 0.0077, *t*-test). DBS also decreases short-term variability in stride length (Fig.4A, *p* = 0.049, *t*-test), toe off angle (Fig. 4C, *p* = 0.029, *t*-test), foot strike angle (Fig.4D, *p* = 0.0029, WSRT), and single-limb support (Fig.4E, *p* = 0.0020, WSRT) as well as long-term variability in single-limb support (Fig. 4E, *p* = 0.042, WSRT).

**Figure 4.**
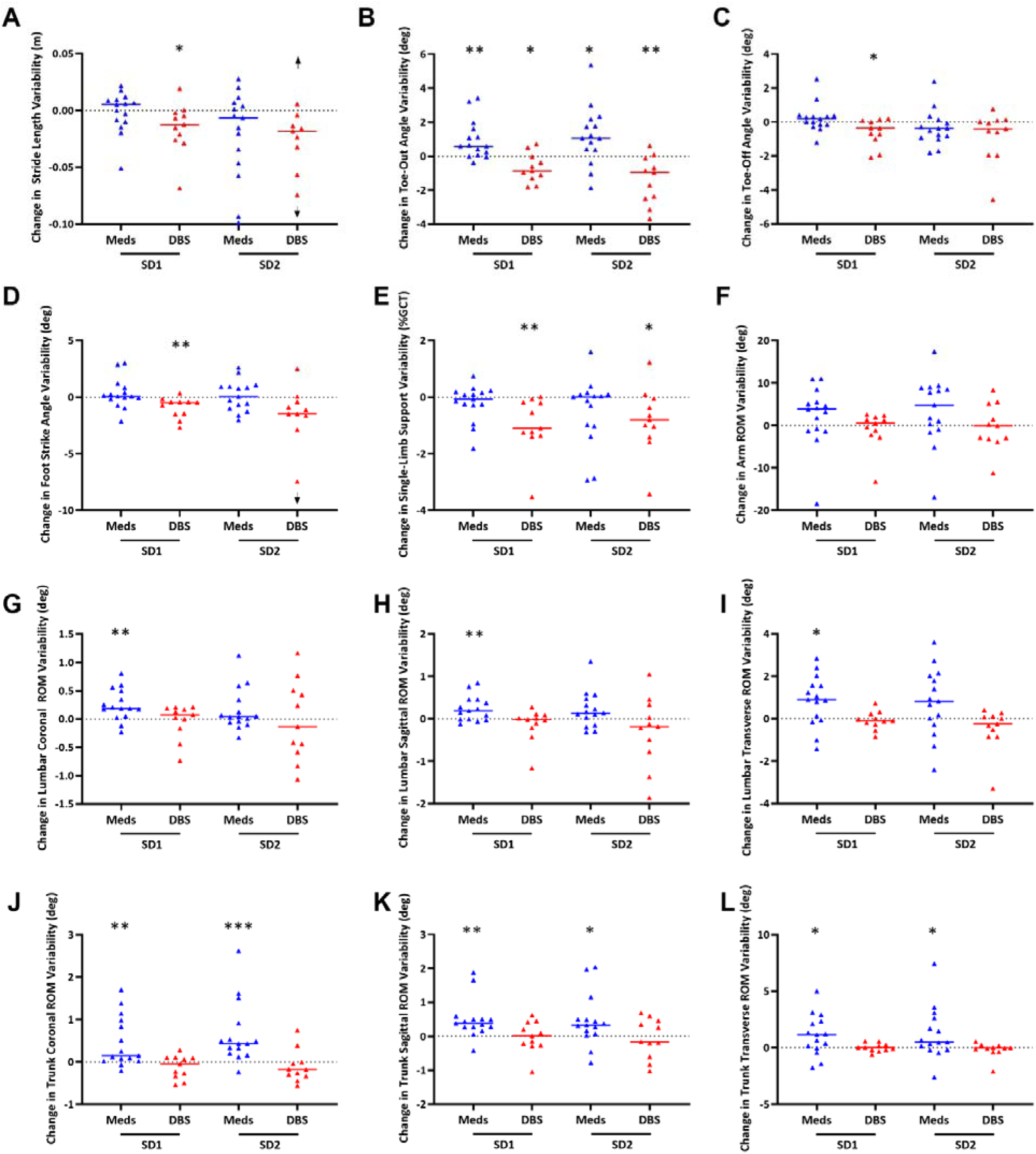
Short-term (SD1) and long-term (SD2) variability changes from STN DBS or medication effect. Abbreviations: W, Wilcoxon signed rank test; pT, paired-T test. * denotes *p* < 0.05, ** denotes *p* < 0.01, and *** denotes p < 0.001. Arrowhead denotes data point out of scale.

Short term variability in lumbar ROM in all planes is significantly increased by medication: coronal (Fig. 4G, *p* = 0.0043, WSRT), sagittal (Fig. 4H, *p* = 0.0074, WSRT), and transverse (Fig. 4I, *p* = 0.026, WSRT). Both SD1 and SD2 for trunk ROM were increased significantly by medication in all planes: coronal (Fig. 4J, *p* = 0.0086, WSRT; *p* = 0.0004, WSRT), sagittal (Fig.4K, *p* = 0.0015, WSRT; *p* = 0.026, WSRT), and transverse (Fig. 4L, *p* = 0.030, *t*-test; *p* = 0.026, WSRT). DBS does not significantly change the variability in any of the measures of body ROM (Fig. 4G-L).

Mean arm ROM variability was increased (in both SD1 and SD2) by medication in 10 out of 15 patients, although the change was not statistically significant. DBS produced no change in arm ROM variability.

## Discussion

Measures of the distribution of values of gait parameters over a period of time (for example the standard deviation of stride length during a 2 minute walk) do not necessarily provide the most accurate measure of gait irregularity, because they contain long-term variability, for example overall slowing due to fatigue over the course of a testing session, which is not necessarily pathological. Short term, step-to-step variability, gives a truer picture of the rhythm disturbance. Poincaré analysis is one way of extracting this information. This technique is commonly employed in quantitative analysis of other physiological phenomena, in particular heart rate variability, and has been used in the analysis of gait (11). Other methods are possible, for example the use of first differences (12).

We found that STN DBS reduces step to step variability in a range of lower limb gait parameters in PD, while antiparkinsonian medication has no significant effect. The lack of effect of medication is at odds with previous literature (1, 3), while the beneficial effects of DBS back the conclusion of the single earlier study (8). Our findings suggest that STN stimulation, but not dopaminergic medication, has access to circuits that control gait rhythm, and that the resulting effect of stimulation on gait is beneficial.

### Cholinergic degeneration underlies some of the abnormalities of gait in PD

PD is typically described in terms of basal ganglial circuit dysfunction arising from the loss of dopaminergic neurons in the substantia nigra pars compacta and their projection to the striatum. While this is the basis for many parkinsonian symptoms, it does not explain the entire syndrome. The pathology includes cholinergic as well as dopaminergic degeneration (13), including neurons of the basal forebrain complex and, of particular relevance here, cholinergic neurons in the pedunculopontine nucleus (PPN) (14-16). The PPN and adjacent cuneate nucleus form the midbrain locomotor region (MLR), which has a major role in coordination of gait and posture. The PPN contains a mixed population of cholinergic, glutamatergic, and GABAergic neurons, and postural instability and gait rhythm disturbances have been linked to its degeneration. Experimentally, postural and gait disturbance can be produced by purely cholinergic PPN lesions without any dopaminergic degeneration (17). The extent of freezing and falling in PD have been found in human postmortem studies to be correlated with the degree of cholinergic PPN neuron loss (17).

The demonstration that levodopa makes no significant difference to the step-to-step variability of a wide range of lower limb parameters of gait in a group of patients with advanced PD, reinforces the conclusion that the maintenance of gait rhythm is not under dopaminergic control. In contrast, acetylcholinesterase inhibitors appear to make gait more regular and stable. Rivastigmine reduces the standard deviation of step time (18), and donepezil has been shown to reduce fall frequency in PD (19).

We have also shown that, unlike dopaminergic medication, DBS of the STN does regularise gait. This suggests that in addition to its well documented benefits to the dopamine dependent symptoms of PD, STN DBS also modulates the behaviour of the cholinergic system. Evidence from PET imaging supports the idea that STN DBS improves Parkinsonian gait by modulating PPN/MLR activity (20). How might it do this? One obvious possibility is that STN stimulation may activate glutamatergic neurons projecting from the STN to the PPN, which then excite cholinergic PPN neurons. However, projections from the STN to PPN are sparse, accounting for some 1% of STN neurons (21). Alternatively, STN DBS might activate glutamatergic STN efferents to the GPi and SNpr, which both project to the PPN and could therefore modulate its activity. However, the GPi and SNpr to PPN connections are both inhibitory. STN stimulation could also be directly activating the cholinergic neurons by retrograde activation of their PPN-STN projections, and we believe this is the more likely explanation for our findings.

### Retrograde activation of the PPN cholinergic arborisation

Cholinergic PPN neurons project very widely, to targets including the substantia nigra (22-28), STN (21, 27, 29), thalamus (27, 28), pallidum (26-28, 30), striatum (27), superior colliculus (26, 31), cerebral cortex (27), and spinal cord. A key anatomical feature of this projection, demonstrated by dual labelling and single neuron tracer studies, is that individual cholinergic PPN neurons project to many of these targets through multiple axon collaterals (26, 32). This raises the possibility that STN stimulation may activate PPN-derived afferents antidromically. Once antidromic APs reach collateral branch points they will propagate down all the collaterals orthodromically, causing widespread activation of PPN targets as if the signals were coming from the PPN itself. Thus STN DBS could act to boost cholinergic PPN activity in a wide range of locations including other parts of the basal ganglia, the MLR itself, and spinal locomotor centres. This is illustrated in figure 5.

**Figure 5:**
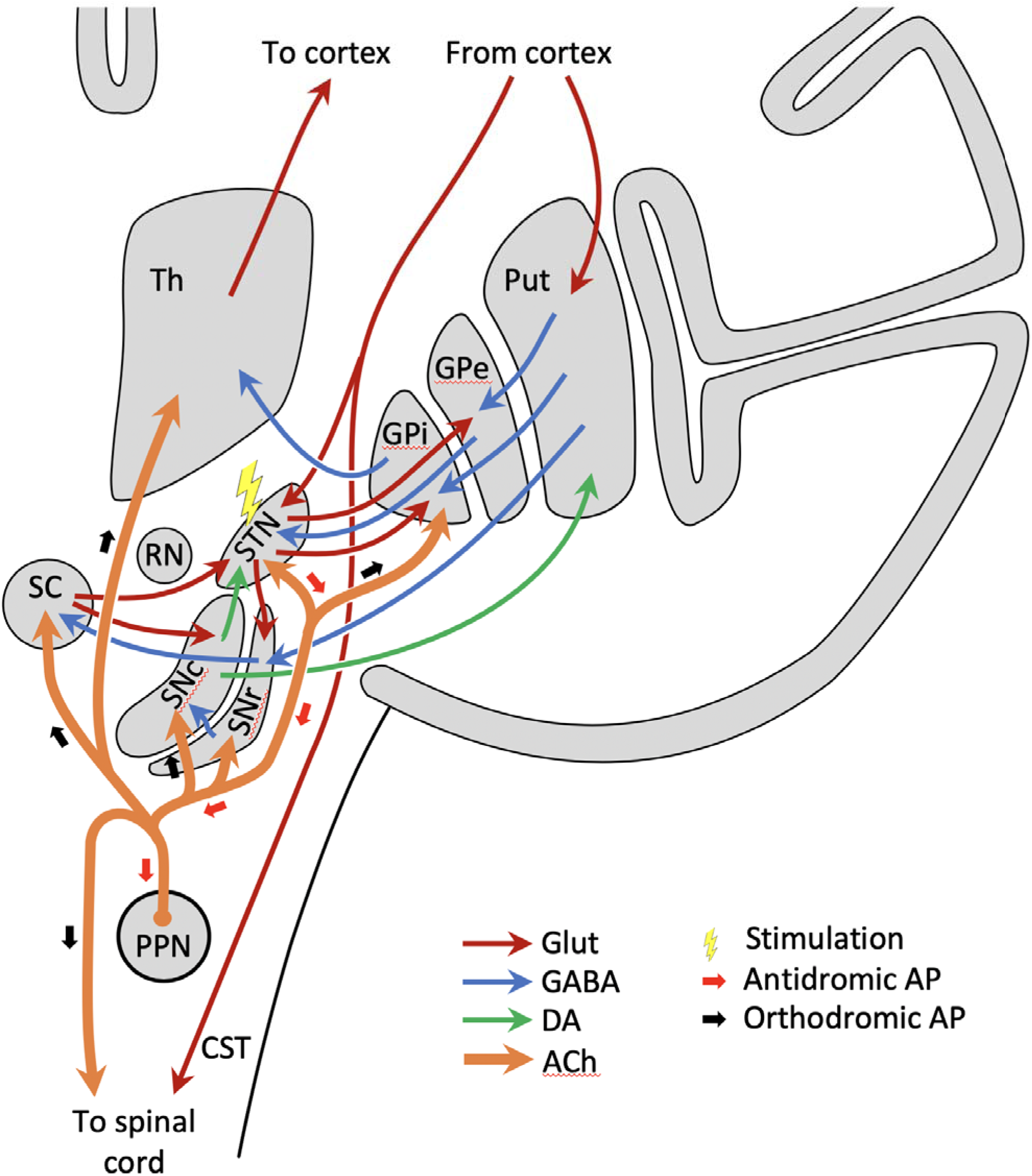
The proposed explanation for the effects shown in the study. Stimulation of the subthalamic nucleus (lightning bolt symbol) results in antidromic action potentials (APs) in afferent cholinergic fibres from the PPN (small red arrows). When these reach branch points they generate orthodromic APs in all of the other collateral branches (small black arrows), activating multiple other PPN projection targets. Th, thalamus; Put, putamen; GPe, globus pallidus externus; GPi, globus pallidus internus; STN, subthalamic nucleus; RN, red nucleus; SNc, substantia nigra pars compacts; SNr, substantia nigra pars reticulata; SC, superior colliculus; PPN, pedunculopontine nucleus; CST, corticospinal tract; Glut, glutamate; GABA, gamma aminobutyric acid; DA, dopamine; ACh, acetyl choline.

We hypothesise that this is the likely mechanism for the dopamine independent improvements in gait rhythm that we have observed with STN DBS here. This hypothesis leads to testable predictions. Firstly, we would expect STN DBS to have cholinergic effects on other nuclei to which the PPN also projects. One such location is the superior colliculus (SC) (31), which receives projections from both the SNpc and the PPN. The SC is a critical structure in the generation of saccades, the rapid conjugate eye movements that shift gaze. Saccades have been studied extensively in PD (33-36) and more specifically in PD patients who have had DBS (37-39). The most commonly used experimental task is the visually guided prosaccade, in which the subject moves their eyes as quickly as possible towards a novel visual target. An important parameter of saccades is their latency, i.e. the time between stimulus presentation and the onset of the saccade. Levodopa produces a mild prolongation of prosaccade latency (PSL) of about 7% (33). In contrast, experiments involving the injection of the cholinergic agonist nicotine into monkey SC *reduced* PSL by 40% (40), while the strong central anticholinergic drug promethazine increased it (41). STN DBS shortens PSL (37, 38, 42, 43), in keeping with a cholinergic, but not a dopaminergic, effect. Furthermore, DBS of the GPi, which also receives cholinergic PPN afferents(30), also shortens PSL to a similar degree (38). In the case of the STN, the DBS effect could be mediated by driving the excitatory projection to the SNpr, which itself sends an inhibitory projection to the SC. However, the route from the GPi is less obvious. Retrograde activation of the PPN arborisation from either location would provide a unifying explanation for the similar effect on saccades.

Secondly, activating the same cholinergic PPN arborisations from a different location should have a comparable effect on gait regularity. In keeping with this, DBS of the GPi, which is carried out for similar symptoms to STN DBS, appears to have a similar beneficial effect on gait (44). To our knowledge however there is no literature examining gait variability in GPi DBS patients in depth, and we intend to investigate this in future work.

Freezing of gait (FoG) can be viewed as an extreme form of rhythm disturbance and it is linked to severe PPN degeneration (17). FoG is reduced by STN DBS in many patients (45-47). Direct stimulation of the PPN itself might be expected to have a more potent effect, and the PPN has been explored by several groups as a target for DBS specifically to treat FoG (48-55). PPN DBS however will not treat dopaminergic deficits and the group of patients to whom it is theoretically best suited – those with freezing but little upper body parkinsonism – is small. This has led some to experiment with dual STN/PPN stimulation (49).

Levodopa can induce FoG in some patients (56). The increased variability in some patients on medication may reflect a mild form of this phenomenon, and the medication group could comprise subgroups that behave differently in this regard. We plan to use the same gait analysis techniques to investigate the response to a levodopa challenge in a larger group of patients in future work.

### Effects on the trunk and upper limbs

The results we observed for movement of the trunk and upper limbs were strikingly different to those seen in the lower limbs. Arm swing ROM was significantly increased by medication, while DBS produced a smaller and non-significant change. Whereas lower limb variability was unaffected by medication and reduced by DBS, upper limb and trunk variability were unaffected by DBS but increased by medication.

There have been many studies in healthy individuals examining to what extent arm swing is passive, with the arm viewed as a mass damper in the form of a pendulum driven by movement at the shoulder (57-59), or active, produced by muscle activity under control of a central pattern generator (60, 61). The likelihood is that it is a combination of the two (62), with a large component of passive motion but added muscle activity, possibly to provide small movement corrections that keep arm swing coordinated with the rest of the body. In PD, arm swing is typically reduced and exhibits an altered phase relationship with lower limb movement. Lower limb bradykinesia in PD may lessen shoulder movement and thus pendulum drive, and increased damping due to parkinsonian rigidity will reduce arm swing amplitude and alter its phase relationship with lower limb movements. The changes in arm swing might also be viewed as evidence for dysfunction in a putative pattern generator for upper limb motion. The fact that medication produces a greater increase in arm ROM than DBS, despite DBS being highly effective in alleviating rigidity, might further suggest that, unlike lower limb gait rhythm, arm swing is under the control of a pattern generator that is dopamine dependent.

However, the trend to increased arm swing variability we observed with medication seems at odds with this as it would suggest that the function of such a pattern generator was more, rather than less, impaired with the addition of exogenous levodopa. A more straightforward interpretation is also possible: that the increased range and variability of arm movements is simply the result of a reduction in damping of arm movement together with higher amplitude and more irregular pendulum drive at the shoulder, due to higher levels of gait variability and consequently higher range and irregularity of trunk movements. This would produce the concomitant increases in amplitude and irregularity of arm swing. The absence of significant change in most of the measures of range and irregularity of arm and trunk movement resulting from DBS may then simply be a consequence of the reduction it produces in lower limb parameter variability.

### Study limitations

DBS patients in this study were on their medication throughout. Some of the effects of DBS on absolute values of gait parameters may have been different in magnitude if testing had been in the medication off state. For example, the relatively small increase in arm ROM with DBS may have been because much of the possible improvement had already been made by medication. Nevertheless, the changes in *variability* are clear and differ not just quantitatively but qualitatively between the groups. We therefore feel that it is unlikely that these are confounded by medication state.

## Data Availability

Anyone interested in the data can email directly to apply for it.

## Acknowledgements

We thank the participants and their families for their endless support with our research work.

## Author’s roles

ZS 1C, 2A and B, 3A and B

SP 1C, 2A and B, 3A and B

BG 1C, 2A and B

TB 2C and 3B

MB 3B

NS 3B

AL 3B

TP 3B

JJF 1A, 1B, 1C, 2A, 2B, 2C and 3A and 3B

CAA 1A, 1B, 1C, 2A, 2B, 2C and 3A and 3B

